# Mathematical Modelling of the Spread of the Coronavirus under Social Restrictions

**DOI:** 10.1101/2020.09.14.20194068

**Authors:** Mo’tassem Al-arydah, Hailay Weldegiorgis Berhe, Khalid Dib, Kalyanasundaram Madhu

**Affiliations:** Department of Mathematics, Khalifa University, P.O.Box: 127788, Abu Dhabi, UAE; Department of mathematics, Mekelle University, Mekelle, Ethiopia

**Keywords:** COVID-19, Coronavirus, mathematical model, variable transmission rate, social restrictions, parameter estimations

## Abstract

**Background:** COVID-19 has affected most countries and declared as pandemic. Most countries have implemented some social restrictions to control it. In this work we will use mathematical modelling to assess the current social restrictions in controlling the spread of the disease.

**Methods:** We formulate a simple susceptible-infectious-recovery (SIR) model to describe the spread of the coronavirus under social restrictions. The transmission rate in this model is considered variable to catch social restrictions impact. We analyze this model, then fit the model to 160 days induced death data in Italy, Iran, USA, Germany, France, India, Spain and China. we estimate some factors that help in understanding not only the spread of the disease but also assess the current social restriction in controlling this disease.

**Results:** We find a formula for the basic reproduction function (*R*(*t*)) and the maximum number of daily infected people. Then estimate the model’s parameters with 95% confidence intervals in these countries. We notice that the model has excellent fit to the disease death data in all considered countries except Iran. The percentage of disease death estimated by the model in Germany and France are 3.8% and 1.2% respectively, which are close to reported percentages values. Finally, we estimate the time, after first reported death, spent under social restrictions to reduce the basic reproduction function (*R*(*t*)) to one unit. The times to do that in Italy, USA, Germany, France, Spain and China are 40, 50, 34,58, 31, and 15 days respectively. However, the Indian social restrictions in the 160 days were not enough to reach *R*(*t*) = 1.

**Conclusion:** The transmission rate is between 0.1035–1.6076 and recovery rate is between 0–0.2456. The disease death rates calculated for Germany and France are more realistic than others with average value 0.0023. Extending the same social restrictions for enough time could control the disease in Italy, USA, Germany, France, India, Spain and China. While, more social restrictions are needed to control the disease in India.

## 1 Introduction

On December 2019, world health organization (WHO), China branch documented various cases of cloud starting pneumonia inside the Wuhan, Hubei, which was named as coronavirus or COVID-19. The coronavirus has affected most countries and taken the lives of many people worldwide. By March 2020, WHO declared that the disease become a pandemic. In fact, it infected more than 21 million with an estimated death rate of 3.62 percent [8]. Many countries have started some social restriction to stop the spread of the virus. Many lives have been saved and economic consequences of the disease have decreased temporary. Also, many scientific community, including mathematicians, have started searching for better way to control the spread of the disease.

Dynamical modeling for the spread of COVID-19 has been performed by many scholars [4, 5, 6]. Some works are adapting time dependent spreading rate [3], which will be adapted in our approach too. A dynamical model with seven compartments was introduced in [1] to describe the transmission of COVID-19 in China. The model was fitted to 30 days local data to estimate the model’s parameters values. A modified SEIR (Susceptible-Exposed-Infected-Recovered) model fitted to short period data in Post-Soviet States to estimate parameters and forecast potential best and worst scenarios for spread of the deadly infection is given in [2]. In this new work we will fit a 160 days collected death data in seven countries to a simple SIR mathematical model and will include the effect of social restriction on parameter values. Finally, because of the similarities between the spread of SARS-CoV and COVID-19, the approach in the SARS work [7] was reviewed to help us in this work.

The work is done in the following way: we will introduce an SIR mathematical model to describe the dynamics of coronavirus under social restrictions. We fit the model to reported death data in some countries to estimate the model’s parameters values. Then we will use the estimated parameter values and the basic reproduction number to investigate the effectiveness of the current social restrictions, in these countries, to control the COVID-19. The work is organized in the following way: In section 2 we introduce the model and define the parameters. In section 3 we discuss the basic reproduction number. In section 4 we describe the data used in this work. In section 5 we discuss the way used to estimate the parameters and the results. Finally, in section 6 we end up the work with summary and conclusion.

## 2 Formulation of the Model

We divide the population into three classes depending on the disease status that are susceptible *S*, infected class *I*, and recovered *R*. Susceptible can acquire infection due to contact with infected class at a time dependent per capita rate *λ*(*t*) (2). Infected recover at natural rate *r*. The disease induced death rate is *μ_d_*. The ODEs that represent the dynamics in this model are

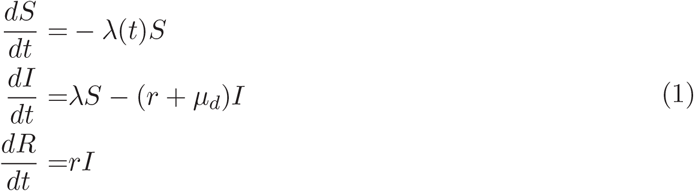

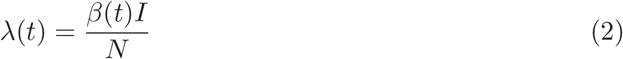

The transmission rate *β*(*t*) is a function of time with *β*(0) = *β*_0_ before social restrictions. Then it decreases with time after applying the social restrictions. The time of applying social restrictions is the time of the first reported death case in this work and the *β* function considered is

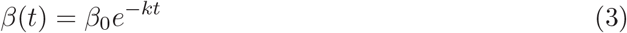

with *k* a new parameter depend on the strength of social restrictions applied (to be estimated). The model’s parameters are described in Table 1. Since 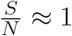, the model is reduced to

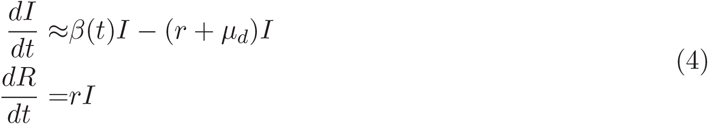

which has the solution

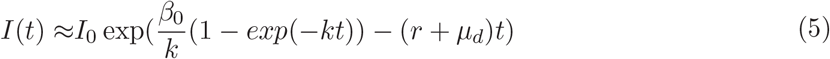

Introducing the cumulative number of infected *I_c_* which can be calculated as follows

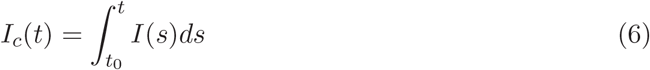

The number of cumulative death from the disease *D_c_* is defined as follows

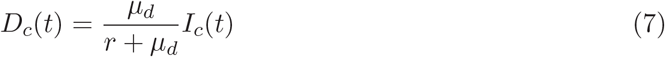

**Table 1:**
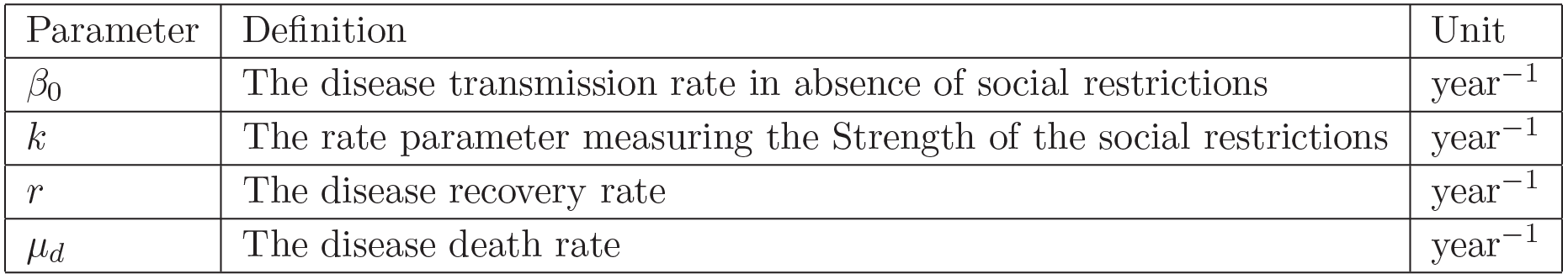
Model’s parameters.

| Parameter | Definition | Unit |
| --- | --- | --- |
| $\beta_0$ | The disease transmission rate in absence of social restrictions | year<sup>-1</sup> |
| $k$ | The rate parameter measuring the Strength of the social restrictions | year<sup>-1</sup> |
| $r$ | The disease recovery rate | year<sup>-1</sup> |
| $\mu_d$ | The disease death rate | year<sup>-1</sup> |

## 3 Basic Reproduction Function

Note that

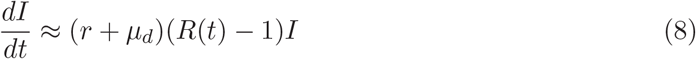

where 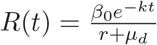. Note that *R*(*t*) is a decreasing function with a maximum value 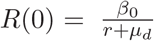. Also *R*(*t*) = 1 iff 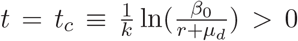. Moreover, *I* is increasing in (0*, t_c_*) and decreasing in (*t_c_, ∞*) with maximum *I* at *t* = *t_c_*. From (5), the maximum number of daily infected is

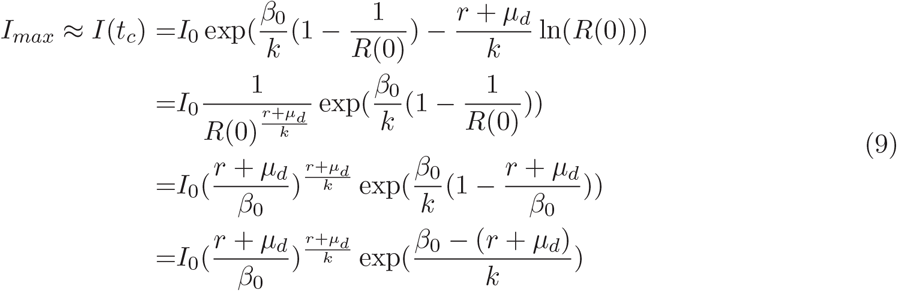

Note that *t_c_* is increasing in *β*_0_ and decreasing in *k, r* and *μ_d_*. The same holds for *I_max_* as *I* is increasing in (0*, t_c_*).

We have found a formula for the time needed to reduce the basic reproduction number to less than one unit and the maximum possible daily infected cases.

## 4 Description of Data

We obtained the updated data of the cumulative number of laboratory confirmed 2019-nCov cases for Italy, USA, Germany, France, India, Spain and China from the WHO (http://www.who.int). The data information includes the cumulative confirmed cases and the cumulative number of deaths from January 21, 2020 till present.

## 5 Parameter Estimations and Analysis

Using the above described data we estimate the model (1) parameters by fitting either the cumulative death function *D_c_* (7) to the reported cumulative death data from the date of the first reported death case to Aug 7. In the following results, we fit the cumulative death function *D_c_* (7) to the reported death cases in the data. The least square method is applied here and implemented in Matlab using the built in function *lsqcurvefit* with the initial parameter guess [*β*_0_*, k, r, μ_d_*] = [0.7, 0.06, 0.03, 0.03], the lower bound *lb* = [0.0001, 0, 0.001, 0.001] and the upper bound *ub* = [2, 0.5, 1, 0.04]. The 95% confidence intervals are calculated using the MATLAB built in function *nlparci*.

### Italy

In Table 2 we have the estimated parameters with their 95% confidence intervals for Italy. Cumulative death graphs for Italy are added to support our results. Figure 1 shows excellent fit between cumulative death from the model and the reported death data for the period Feb 21-Aug 7. Moreover, the Figure 2 shows how the reproduction function changes with time when applying social restrictions to control the disease. The figure shows that after the first reported death case, it took the government in Italy 40 days to reduce *R*(*t*) to one unit.

**Table 2:** Parameter values for Italy using reported cases Feb 21-Aug 7, 2020.

| parameter | value | unit | 95% confidence interval |
| --- | --- | --- | --- |
| $\beta_0$ | 0.7227 | year<sup>-1</sup> | (0.6457, 0.7996) |
| $k$ | 0.0702 | year<sup>-1</sup> | (0.0662, 0.0741) |
| $r$ | 0.0332 | year<sup>-1</sup> | (0.0254, 0.0410) |
| $\mu_d$ | 0.0135 | year<sup>-1</sup> | (0.0047, 0.0223) |

**Figure 1:**
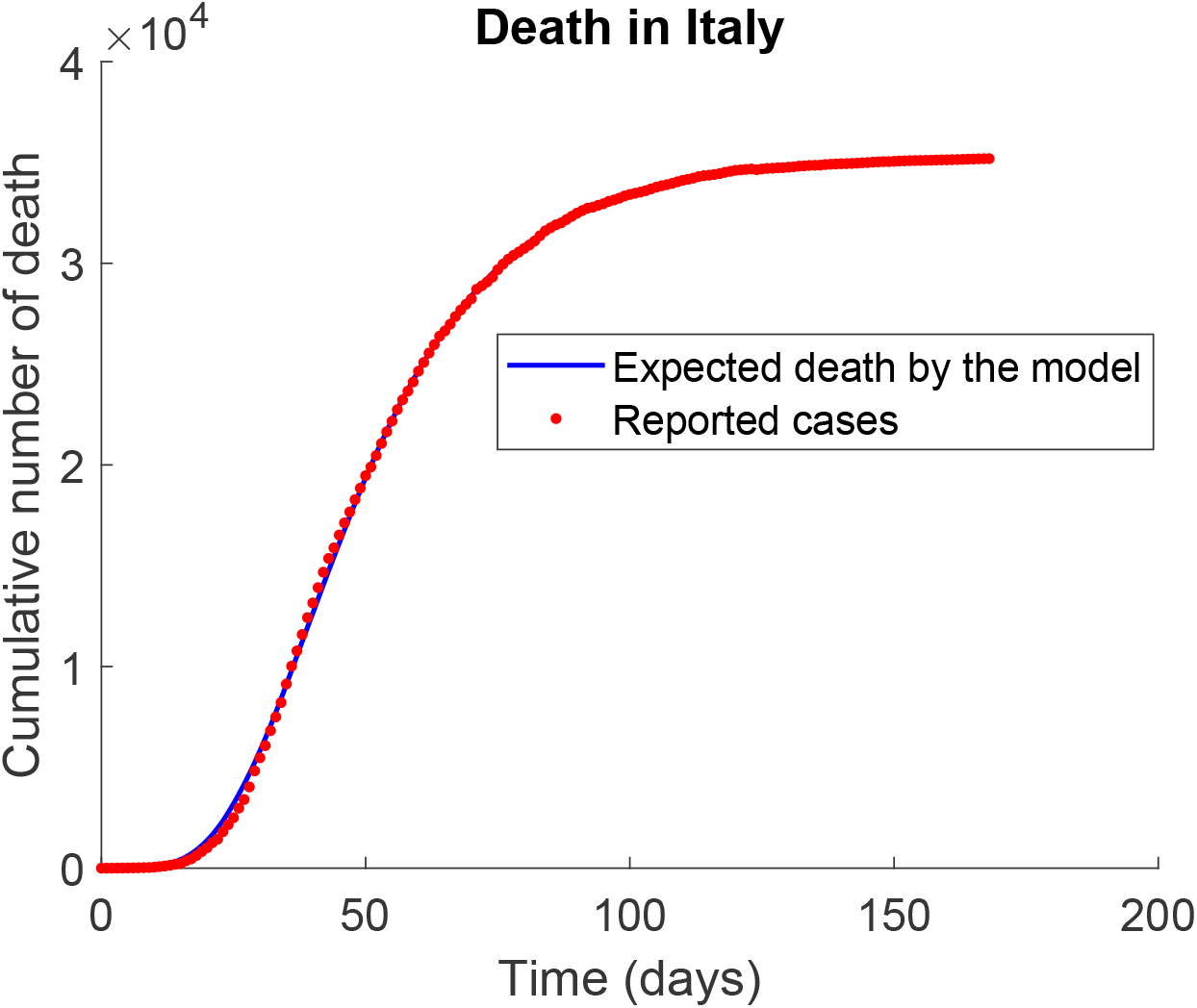
Expected and reported cumulative death cases in Italy.

**Figure 2:**
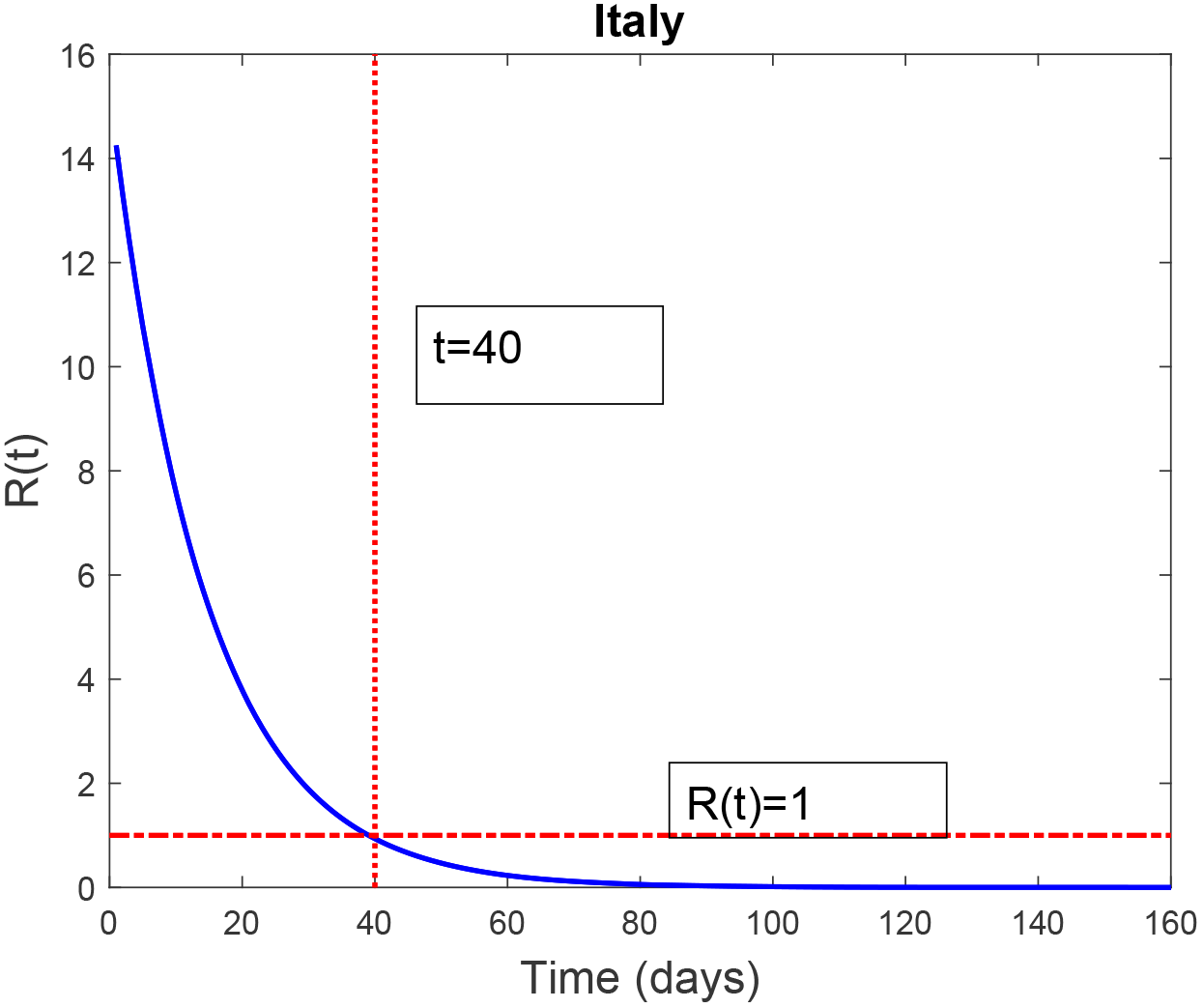
Basic reproduction function *R*(*t*) in Italy.

### USA

In Table 3 we have estimated parameters with their 95% confidence intervals obtained for the USA. The confidence interval for the positive parameter *μ_d_* should be ignored here as it has negative lower bound. Cumulative death graphs for USA are added to support our results.

Figure 3 shows excellent fit between the cumulative death function and the reported death data Feb 21-Aug 7. Moreover, Figure 4 shows how the reproduction function changes with time when applying social restrictions to control the disease. The figure shows that after the first reported death case, it took the government in the USA 50 days to reduce *R*(*t*) to one unit.

**Table 3:**
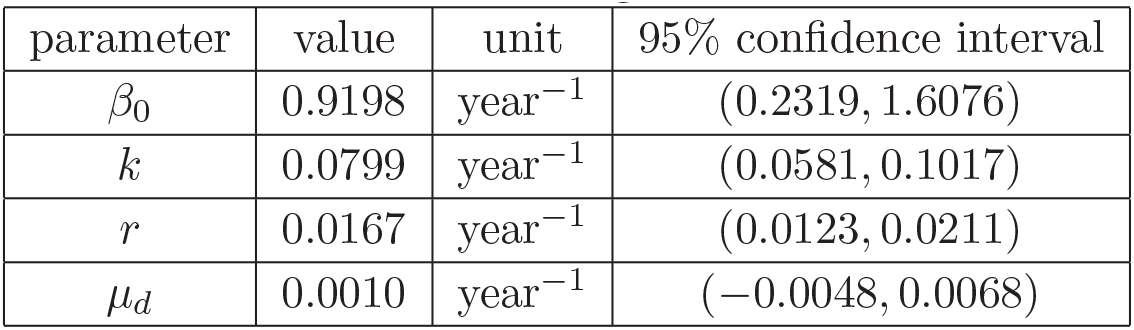
Parameter values for USA using reported cases Feb 29-Aug 7, 2020.

**Figure 3:**
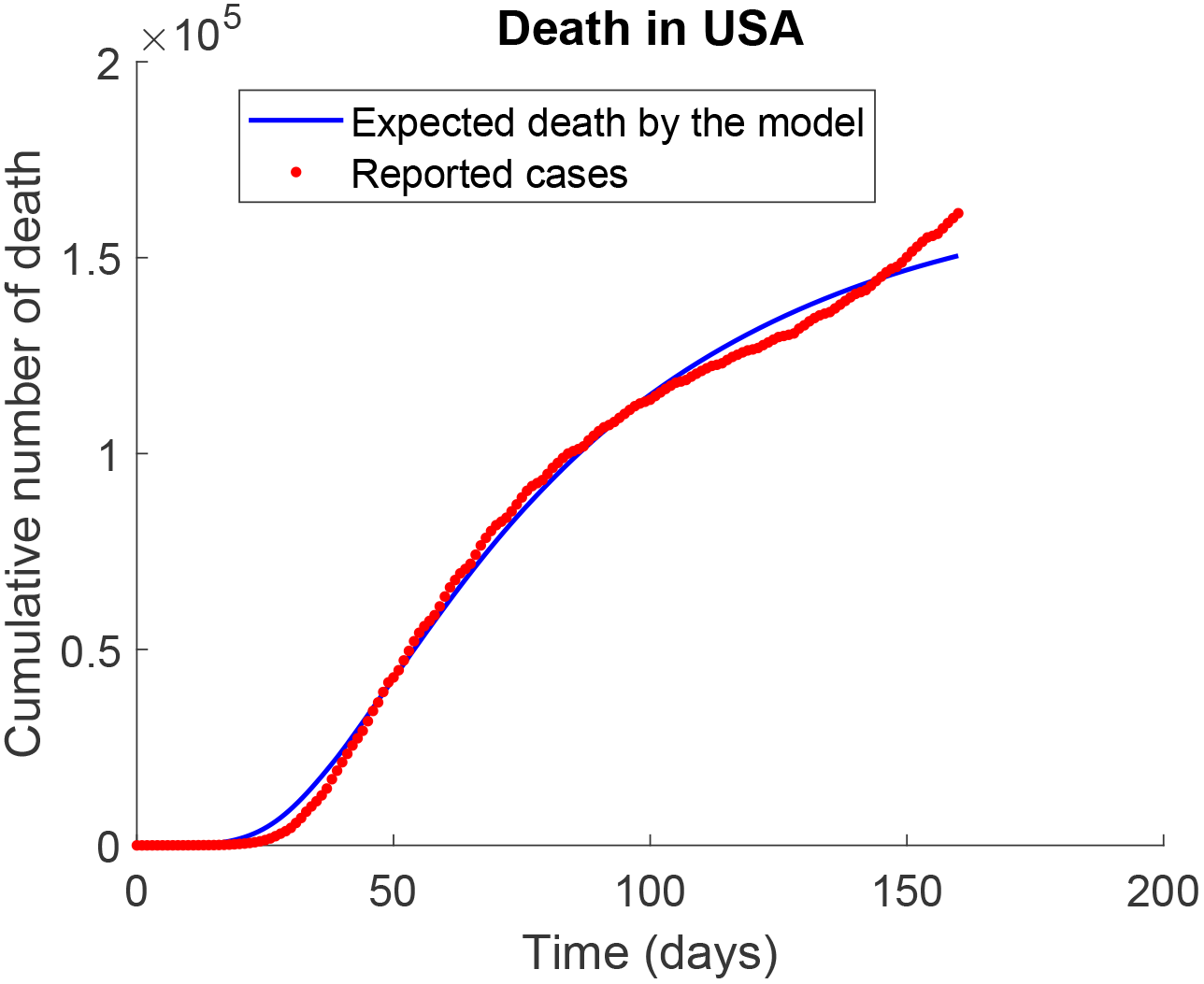
Expected and reported cumulative death cases in the USA.

**Figure 4:**
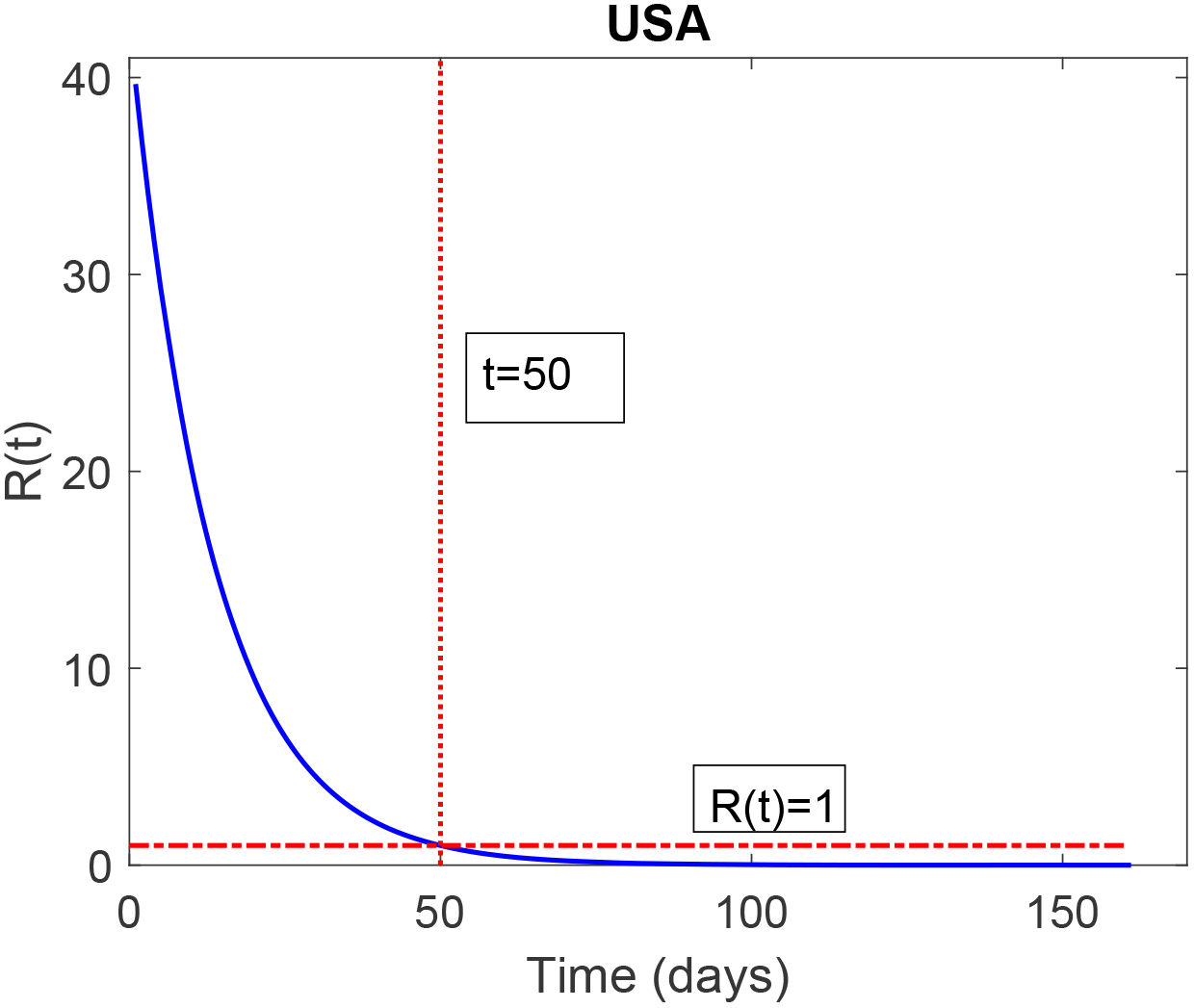
Basic reproduction function *R*(*t*) in the USA.

### Germany

In Table 4 we have estimated parameters values with their 95% confidence intervals for Germany. The confidence interval for the positive parameter *μ_d_* has negative part and should be ignored. Cumulative death graphs for Germany are added to support our results. Figure 5 shows excellent fit between cumulative death function and the reported death data March 9-Aug 7. Moreover, Figure 6 shows how the reproduction function changes with time when applying social restrictions to control the disease. The figure shows that, after the first observed death, it took 34 days in Germany to reduce *R*(*t*) to one unit. Note that percentage of corona death defined by 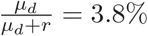 which close to that reported in Germany.

**Table 4:** Parameter values for Germany using reported cases March 9-Aug 7, 2020.

| parameter | value | unit | 95% confidence interval |
| --- | --- | --- | --- |
| $\beta_0$ | 0.8356 | year<sup>-1</sup> | (0.7029, 0.9683) |
| $k$ | 0.0783 | year<sup>-1</sup> | (0.0716, 0.0851) |
| $r$ | 0.0558 | year<sup>-1</sup> | (0.0543, 0.0574) |
| $\mu_d$ | 0.0022 | year<sup>-1</sup> | (-0.0000, 0.0045) |

**Figure 5:**
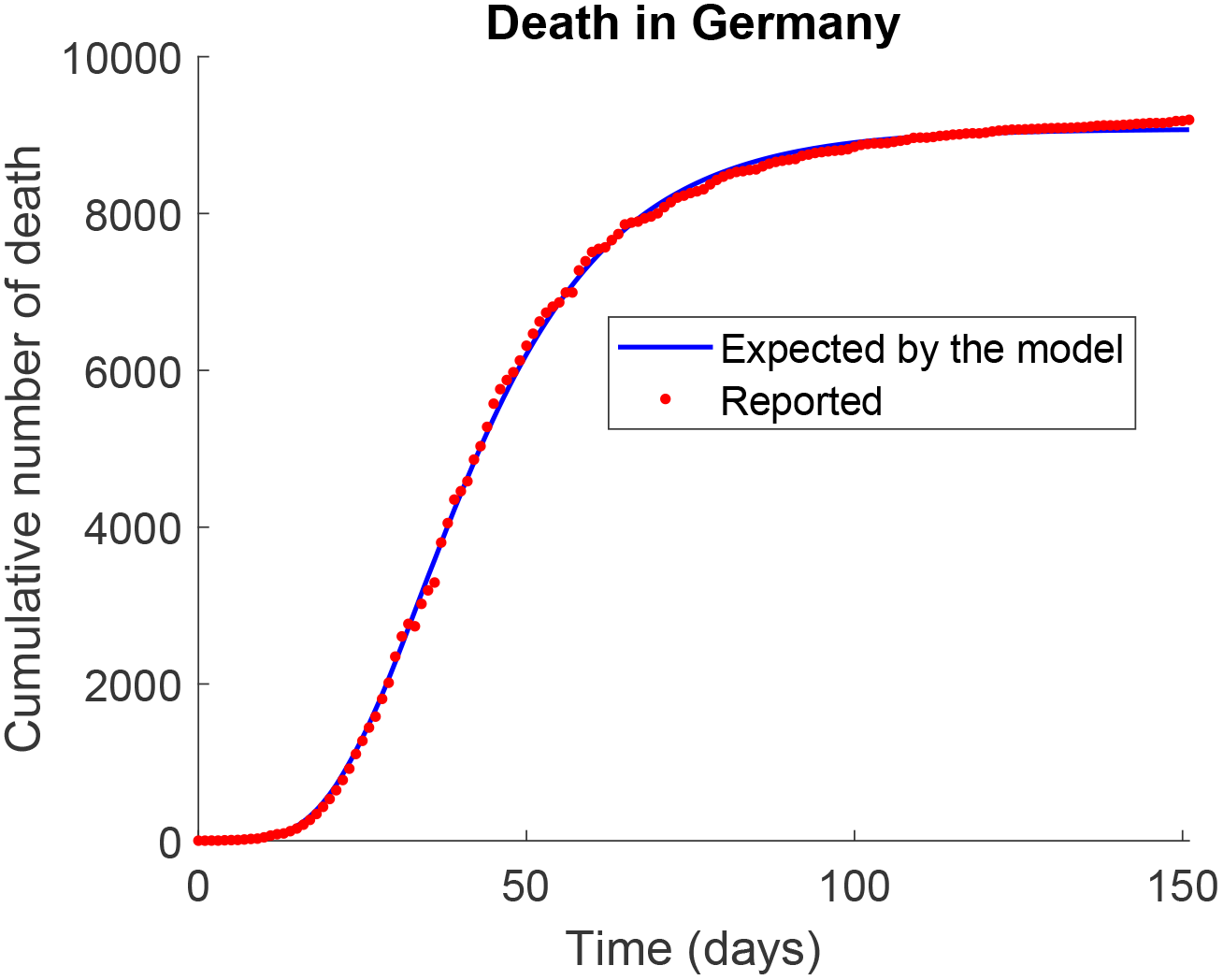
Expected and reported cumulative death cases in Germany.

### France

In Table 5 we have parameter estimates with their 95% confidence intervals obtained for France. The confidence interval for *μ_d_* > 0 should be ignored. Cumulative death graphs for France are added to support our results. Figure 7 shows excellent fit between cumulative death function and the reported death data Feb 15-Aug 7. Moreover, Figure 8 shows how the reproduction function changes with time when applying social restrictions to control the disease. In France, it took 58 days, after the first death, to reduce *R*(*t*) to one unit. Note that percentage death to be 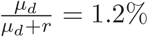, which is close to the reported percentage.

**Figure 6:**
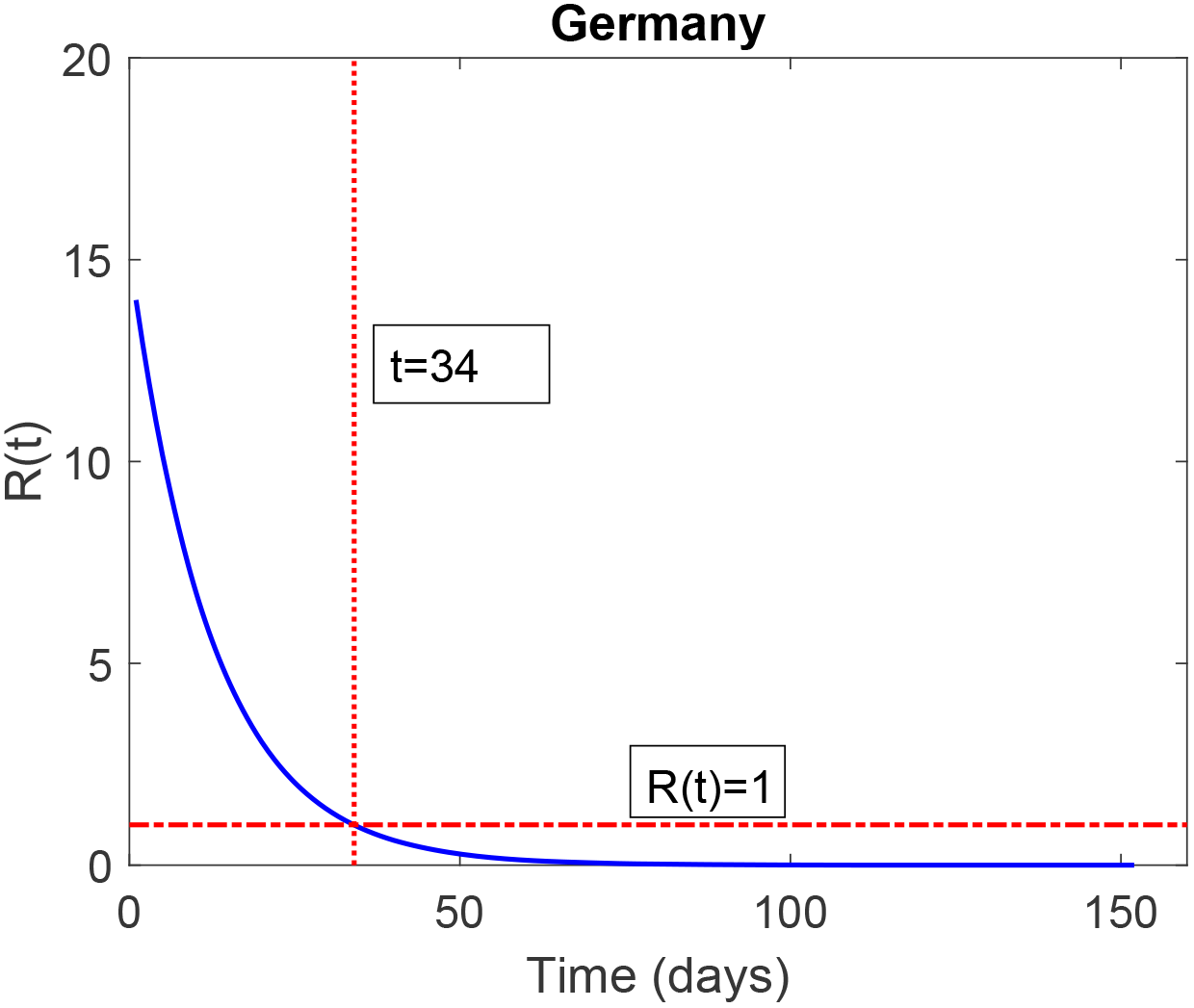
Expected and reported cumulative death cases in Germany.

**Table 5:** Parameter values for France using reported cases Feb 15-Aug 7, 2020.

| parameter | value | unit | 95% confidence interval |
| --- | --- | --- | --- |
| $\beta_0$ | 0.6619 | year<sup>-1</sup> | (0.5185, 0.8052) |
| $k$ | 0.0239 | year<sup>-1</sup> | (0.0110, 0.0368) |
| $r$ | 0.1683 | year<sup>-1</sup> | (0.0909, 0.2456) |
| $\mu_d$ | 0.0024 | year<sup>-1</sup> | (-0.0065, 0.0112) |

### India

In Table 6 we have parameter estimates with their 95% confidence intervals obtained for India. *r* and *μ_d_* confidence intervals should be ignored. Cumulative death graphs for India are added to support our results. Figure 9 shows excellent fit between cumulative death function and the reported death data Mar 11-Aug 7. Figure 10 shows that the social restrictions in India in this period were not enough to reduce *R*(*t*) to one unit.

### Spain

In Table 7 we have parameter estimates with their 95% confidence intervals obtained for Spain. The confidence interval for *μ_d_* should be ignored here. Cumulative death graphs for Spain are added to support our results. Figure 11 shows excellent fit between cumulative death function and the reported death data Mar 3-Aug 7. Moreover, Figure 12 shows how the reproduction function changes with time when applying social restrictions to control the disease. After the first death in Spain, the social restrictions took 31 days to reduce *R*(*t*) to one unit.

**Figure 7:**
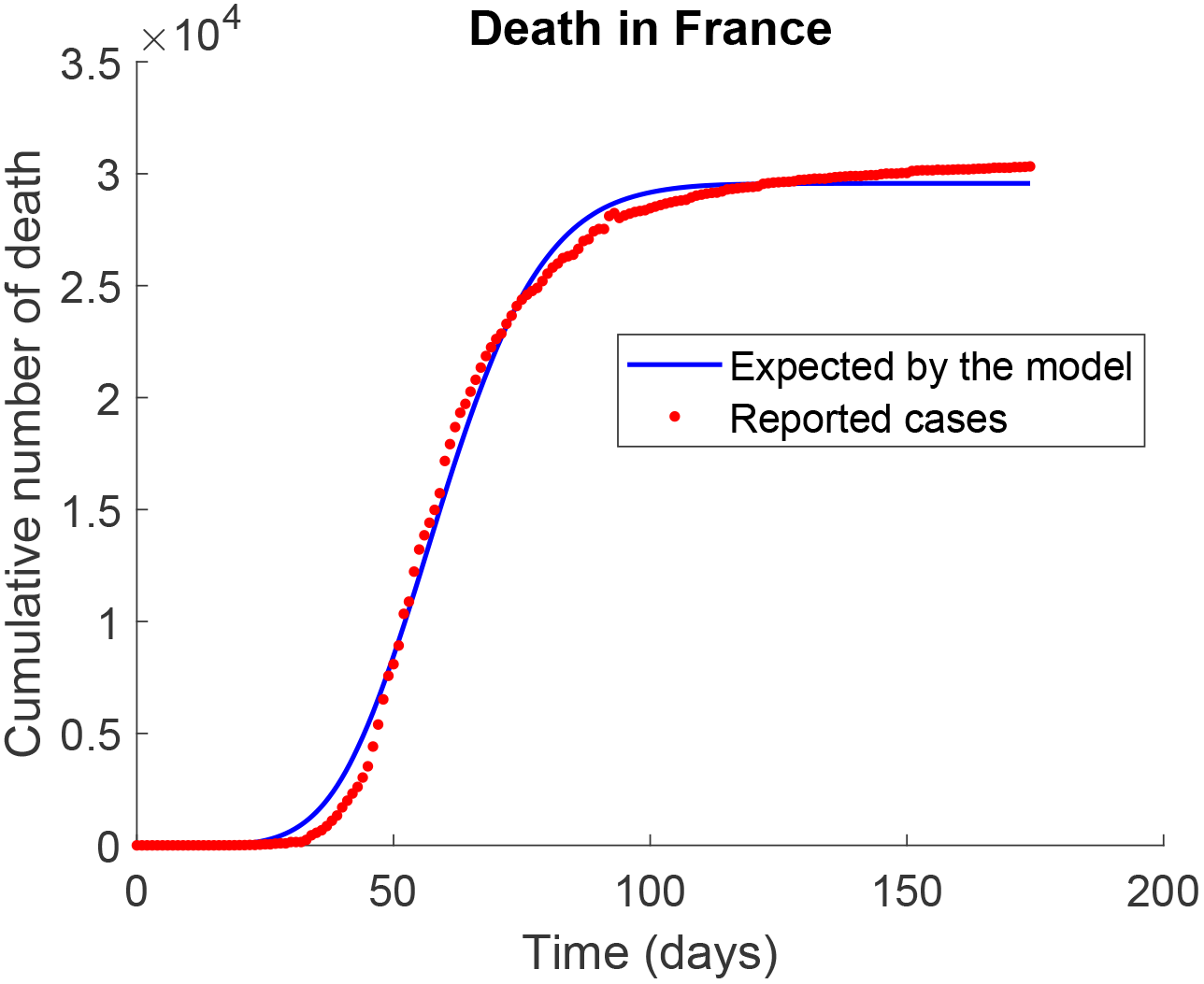
Expected and reported cumulative death cases in France.

**Figure 8:**
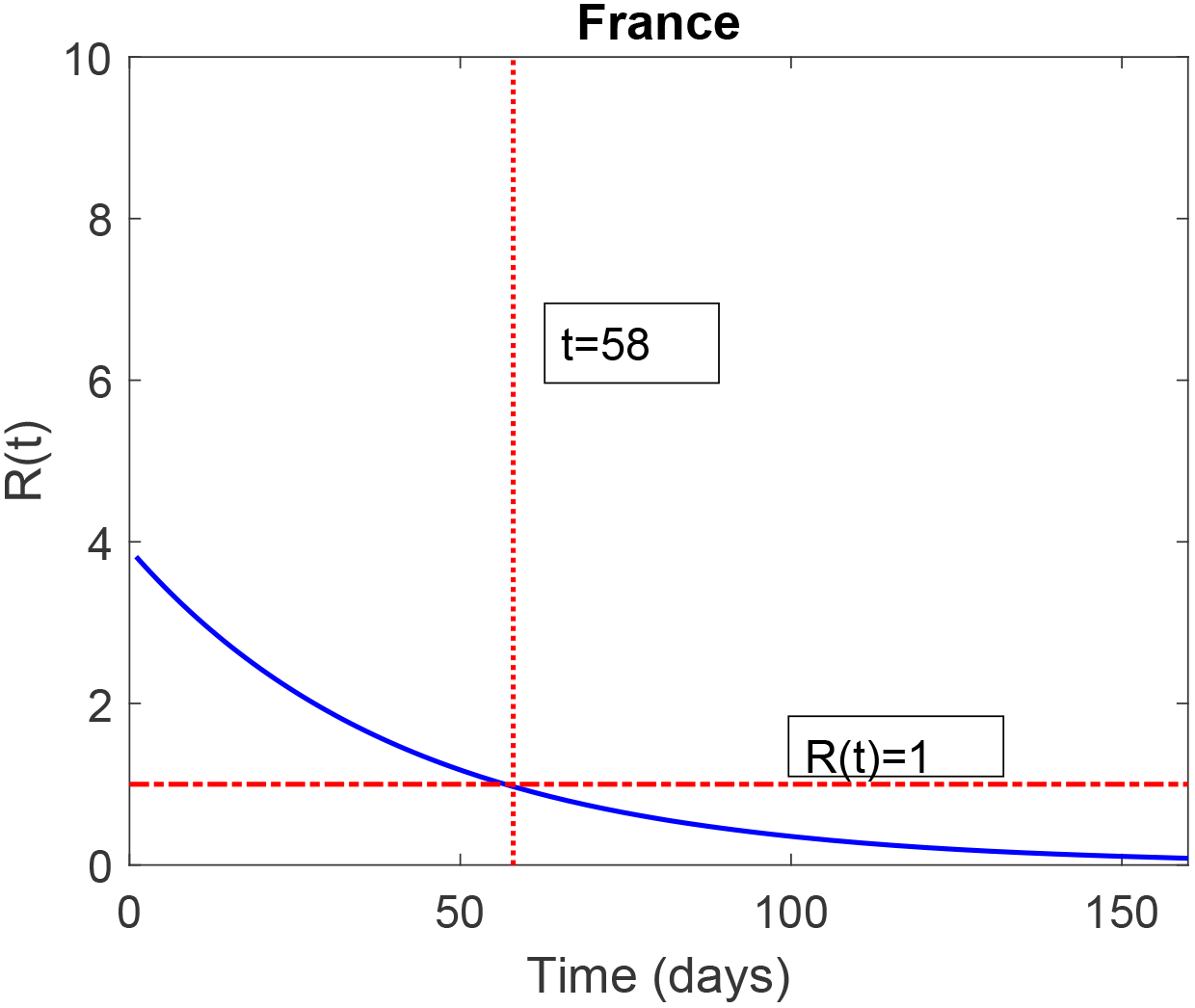
Expected and reported cumulative death cases in France.

**Figure 9:**
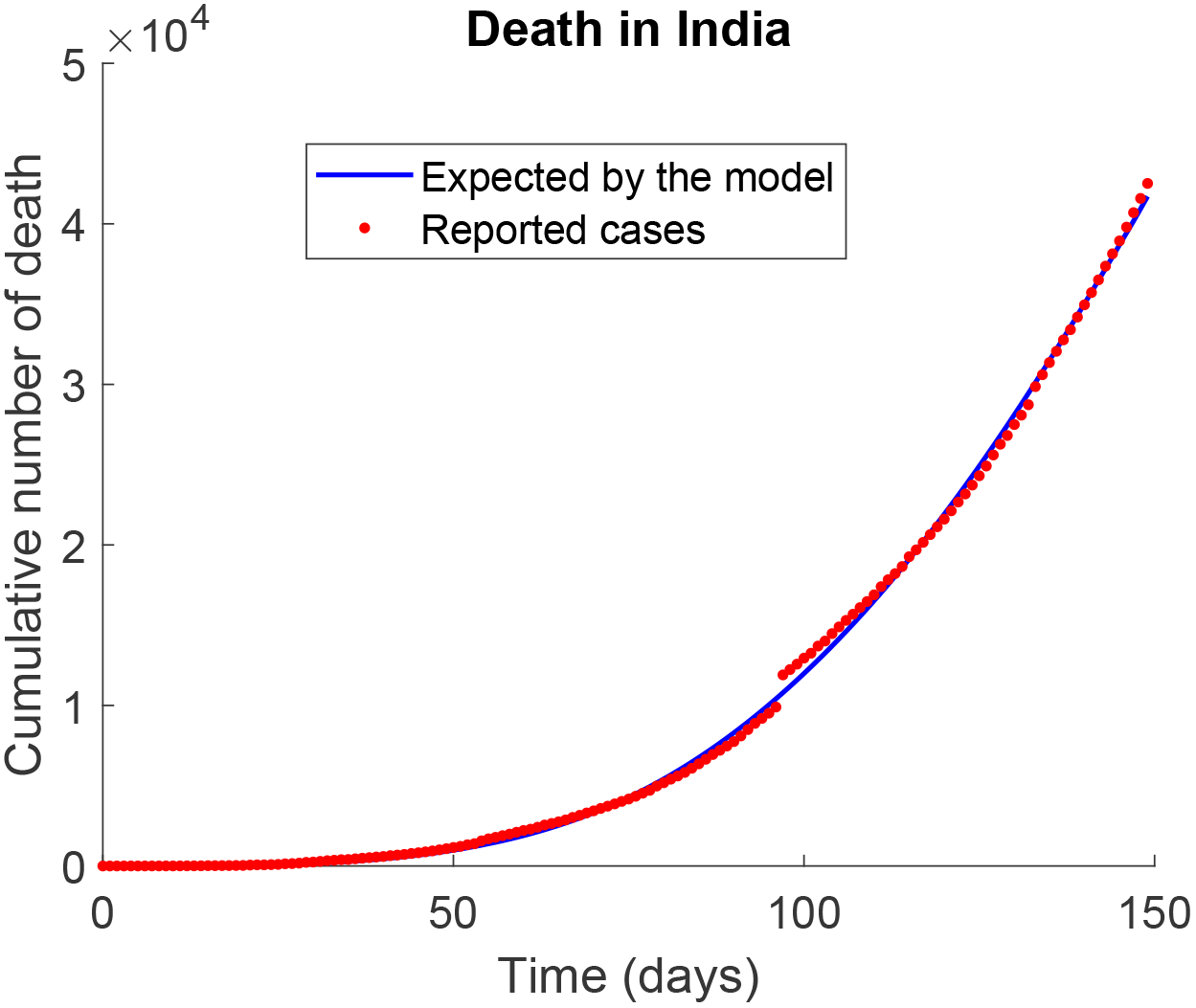
Expected and reported cumulative death cases in India.

**Figure 10:**
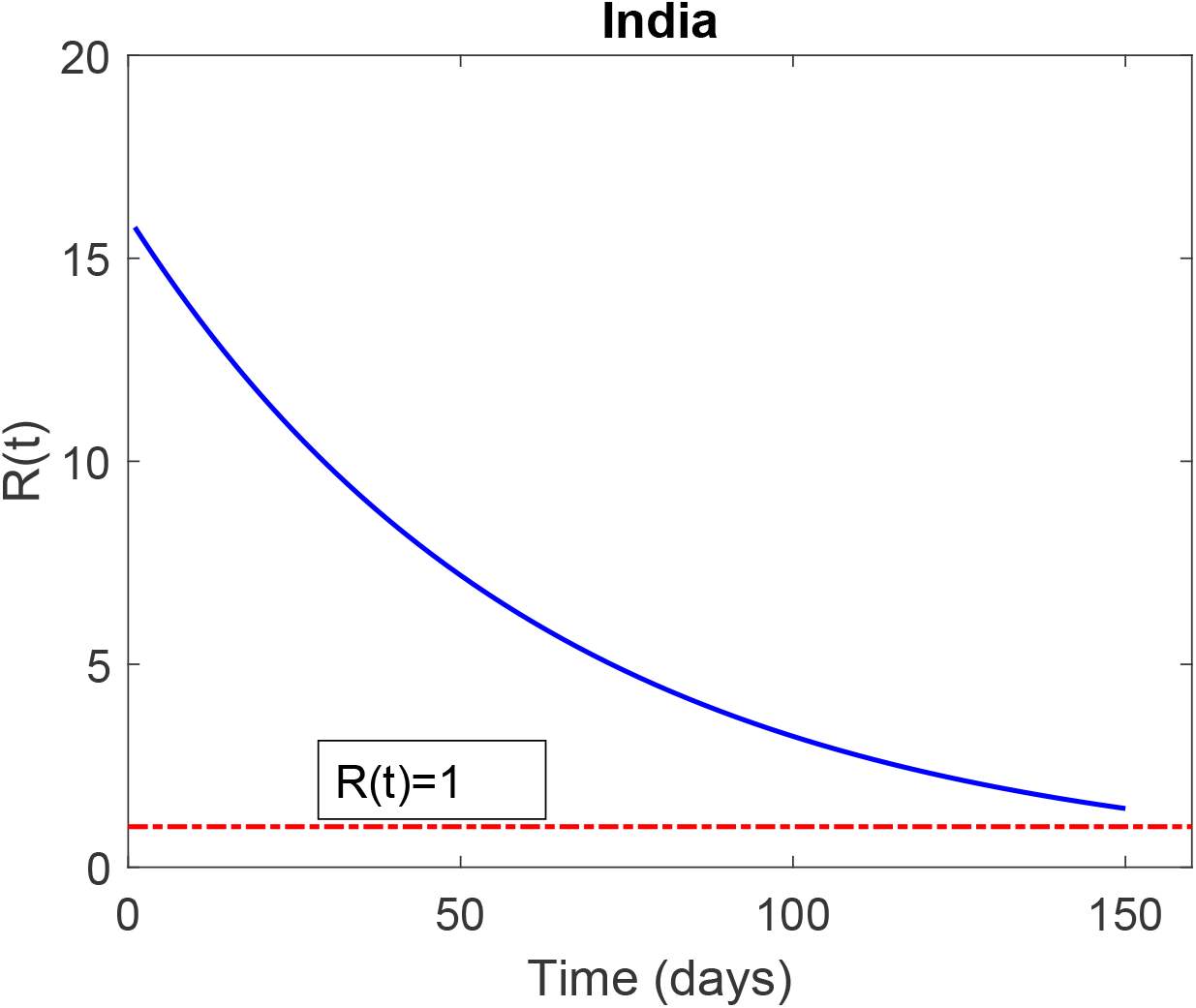
Expected and reported cumulative death cases in India.

**Figure 11:**
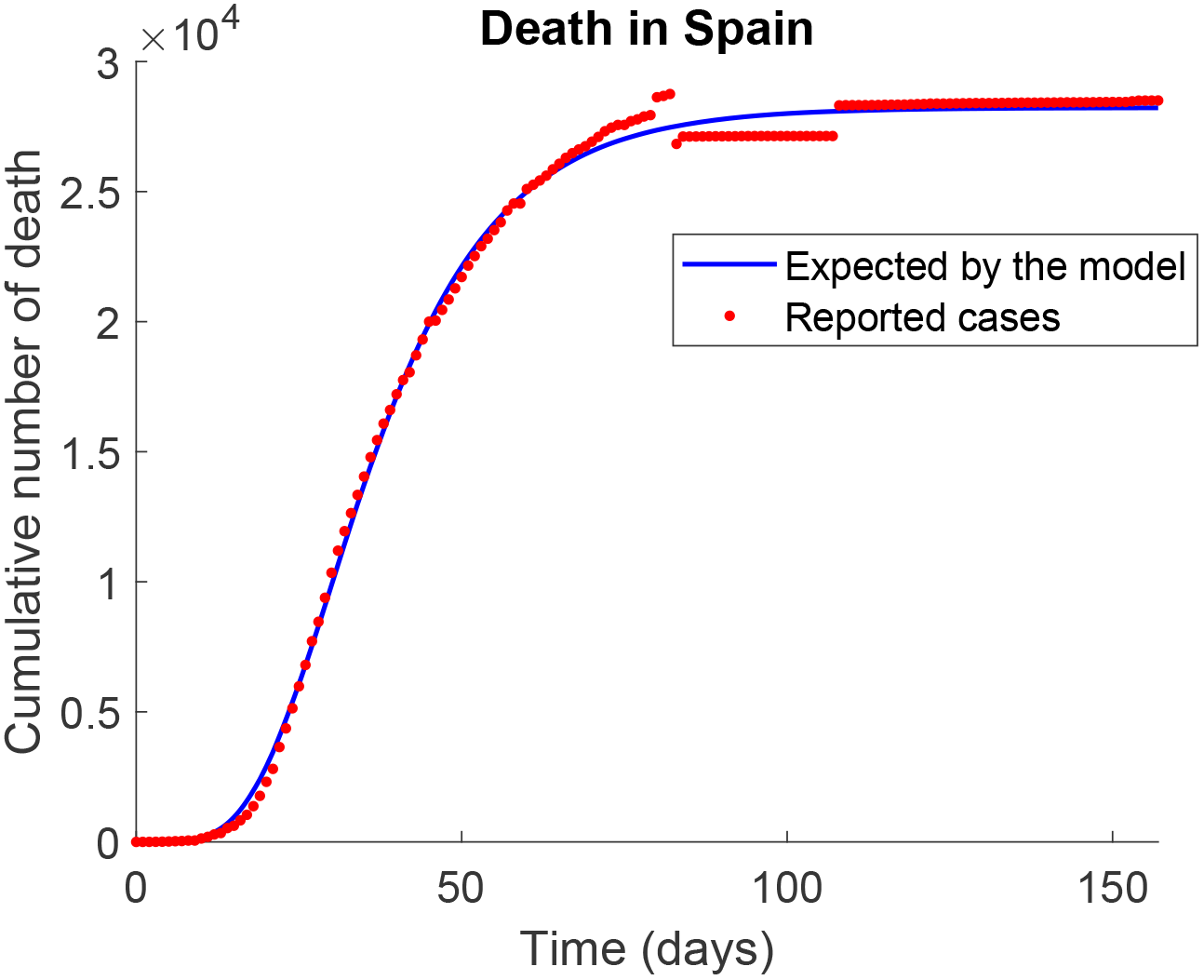
Expected and reported cumulative death cases in Spain.

**Figure 12:**
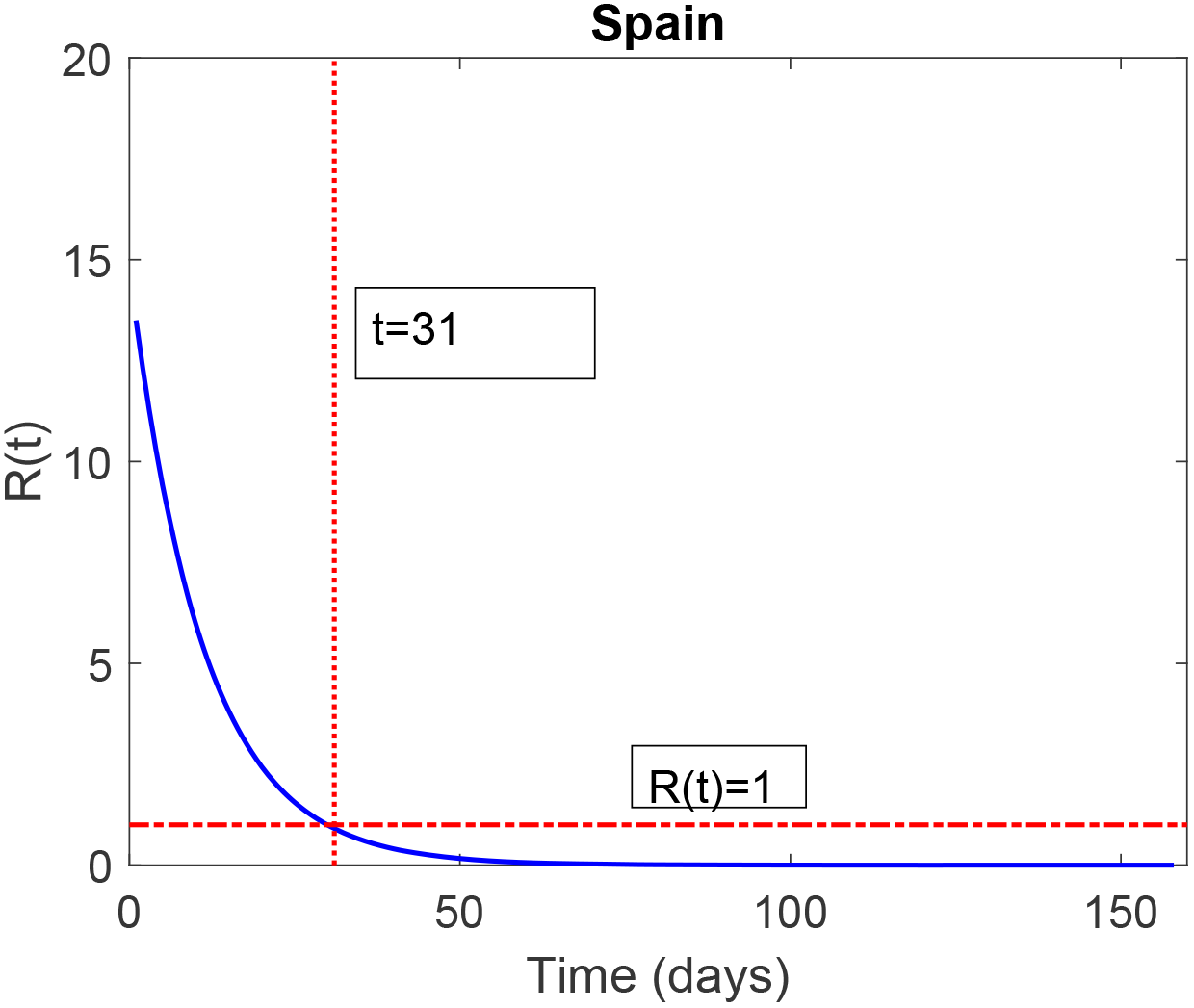
Expected and reported cumulative death cases in Spain.

**Table 6:** Parameter values for India using reported cases Mar 11-Aug 7, 2020.

| parameter | value | unit | 95% confidence interval |
| --- | --- | --- | --- |
| $\beta_0$ | .1425 | year<sup>-1</sup> | (0.1035, 0.1815) |
| $k$ | 0.0176 | year<sup>-1</sup> | (0.0087, 0.0265) |
| $r$ | 0.0010 | year<sup>-1</sup> | (0, 0.0027) |
| $\mu_d$ | 0.0027 | year<sup>-1</sup> | (-0.0106, 0.0160) |

**Table 7:** Parameter values for Spain using reported cases Mar 3-Aug 7, 2020.

| parameter | value | unit | 95% confidence interval |
| --- | --- | --- | --- |
| $\beta_0$ | 0.9838 | year<sup>-1</sup> | (0.6711, 1.2966) |
| $k$ | 0.0897 | year<sup>-1</sup> | (0.0746, 0.1048) |
| $r$ | 0.0528 | year<sup>-1</sup> | (0.0273, 0.0782) |
| $\mu_d$ | 0.0143 | year<sup>-1</sup> | (-0.0153, 0.0440) |

### China

In Table 8 we have parameter estimates with their 95% confidence intervals obtained for China. All confidence intervals here should be ignored. Cumulative death graphs for China are added to support our results. Figure 13 shows excellent fit between cumulative death function and the reported death data Jan 22-Aug 7. Moreover, Figure 14 shows how the reproduction function changes with time when applying social restrictions to control the disease. After the first reported death in China, social restrictions applied reduces *R*(*t*) to one unit in 15 days.

**Table 8:** Parameter values for China using reported cases Jan 22-Aug 7, 2020.

| parameter | value | unit | 95% confidence interval |
| --- | --- | --- | --- |
| $\beta_0$ | 0.7240 | year<sup>-1</sup> | (-1.1520, 2.6000) |
| $k$ | 0.2242 | year<sup>-1</sup> | (-0.0262, 0.4747) |
| $r$ | 0.0164 | year<sup>-1</sup> | (-0.0228, 0.0556) |
| $\mu_d$ | 0.0080 | year<sup>-1</sup> | (-0.0314, 0.0474) |

The fitting for Iran data is poor and dropped from the results presented here. Although, the death data reported for both China and Spain had unreasonable jumps, we ignored these unreasonable reported data in our work and considered the data as correct data.

We have estimated the parameters for the SIR model with variable transmission rate in Italy, USA, Germany, France, India, Spain and China. Also, we have found the 95% confidence intervals for these parameters in each country. Moreover, we have calculated the *I_max_* and *t_c_* estimated by the model and the maximum daily infections reported by data and the day when reported.

**Figure 13:**
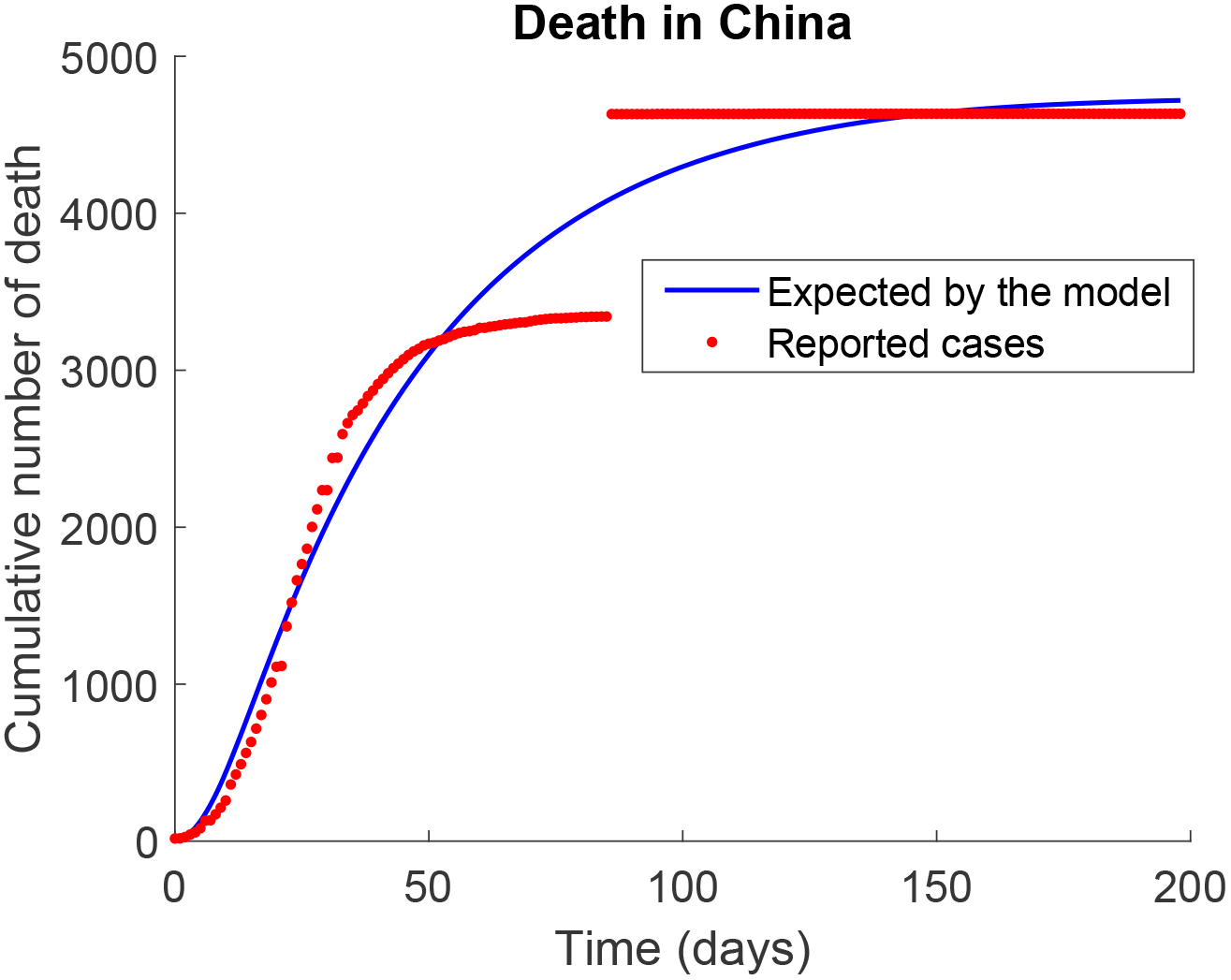
Expected and reported cumulative death cases in Spain.

**Figure 14:**
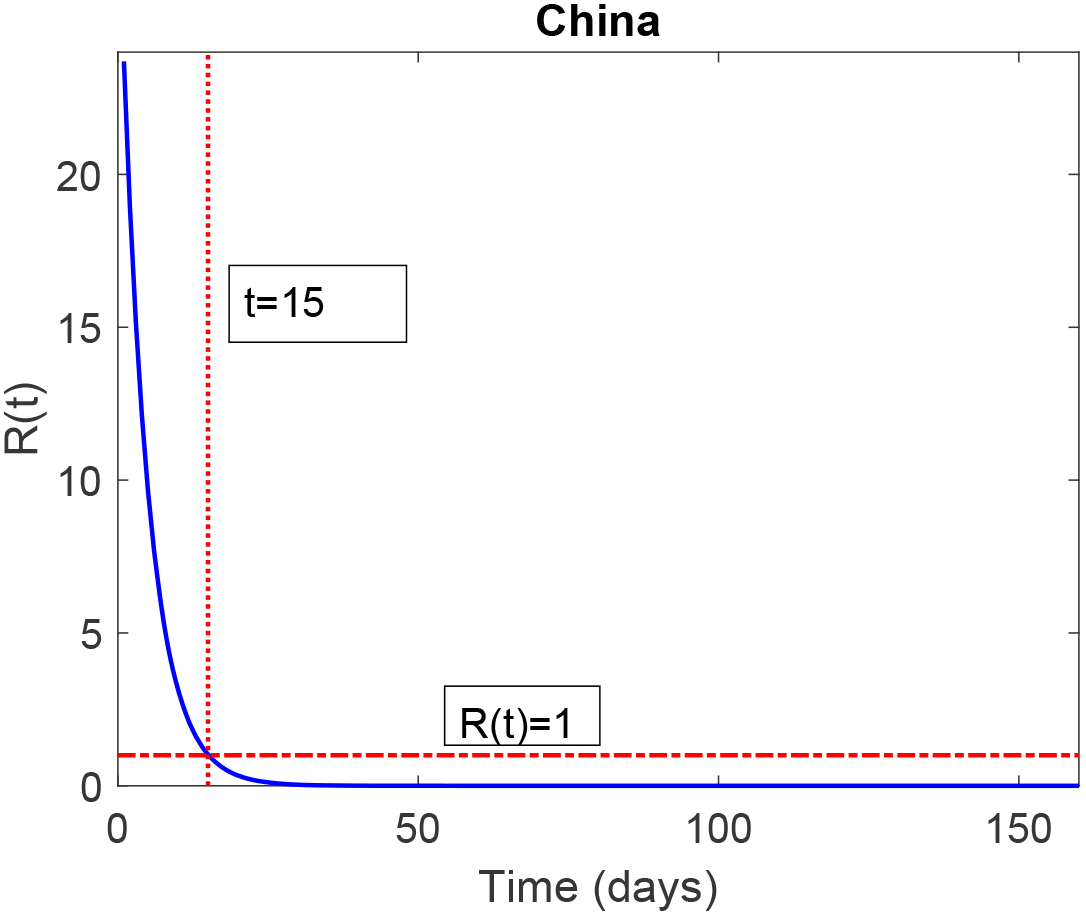
Expected and reported cumulative death cases in Spain.

## 6 Summary and Conclusion

We have introduced a simple SIR model to describe the spread of the coronavirus under social restriction. The rate of the transmission of the disease is considered time dependent due to countries lock down and quarantine. The model was fitted to induced death data and the parameter values are estimated with 95% confidence intervals in Italy, USA, Germany, France, India, Spain and China. Combining the confidence intervals for the parameters *β*_0_, *k* and *r* for all the countries, we ended up with the global intervals listed in Table 9.

**Table 9:** Interval for parameter values regardless of country but excluding China.

| Parameter | Interval for the value |
| --- | --- |
| $\beta_0$ | (0.1035, 1.6076) |
| $k$ | (0.0087, 0.1048) |
| $r$ | (0, 0.2456) |

The model shows that the social restrictions in China were the most effective as it reduced *R*(*t*) to one unit within 15 days after the first reported death case. However, the Indian social restrictions after the first death were not enough to reach *R*(*t*) = 1 in 160 day. The times, after the first death case, spent to reduce *R*(*t*) to one unit, which could lead to controlling the coronavirus, are summarized in Table 10. Although the fitting was excellent in most countries, the fitting in Germany and France are more realistic as the percentages of induced death estimated by the model in the two countries are 3.8% and 1.2% respectively which are close to the reported worldwide value 3.6%.

**Table 10:** Time, after the first reported death, needed to reduce *R*(*t*) to one Country Time (days)

| Country | Time (days) |
| --- | --- |
| Italy | 40 |
| USA | 50 |
| Germany | 34 |
| France | 58 |
| India | - |
| Spain | 31 |
| China | 15 |

It should be mentioned that the *I_c_*(*t*) has been fitted to infected cases too. Although we got good fit in Italy and China (only), the parameter values found are different from the parameter values estimated using induced death data. As the authors believe that death data could be more accurate than infected reported data, we drop these results from this work.

The work has some limitations: In some countries we couldn’t estimate the confidence interval for the disease death rate. The parameters values estimated here are average values and vary within the estimated confidence interval, readers should use them carefully. For future work, more countries and longer period data will be used to check the validity of current results.

## Data Availability

Data are available

https://covid19.who.int/

## References

[1] Jiwei J, Jian D, Siyu L, Guidong L, Jingzhi L, Ben D, Guoqing W and Ran Z. Modelling the control of COVID-19: impact of policy interventions and meteorological factors, Electronic Journal of Differential Equations, 2020(23):1–24.

[2] Issanov A, Amanbek Y, Abbay A, Adambekov S, Aljofan M, Kashkynbayev A, Gaipov A. COVID-19 Outbreak in Post-Soviet States: Modeling the Best and Worst Possible Scenarios. Electron J Gen Med. 2020;17(17):em256. https://doi.org/10.29333/ejgm/8346

[3] Cakir Z, Savas HB. A Mathematical Modelling Approach in the Spread of the Novel 2019 Coronavirus SARS-CoV-2 (COVID-19) Pandemic. Electron J Gen Med. 2020;17(4):em205. https://doi.org/10.29333/ejgm/7861

[4] Li Y, Wang B, Peng R, Zhou C, Zhan Y, Liu Z, et al. Mathematical Modeling and Epidemic Prediction of COVID-19 and Its Significance to Epidemic Prevention and Control Measures. Ann Infect Dis Epidemiol. 2020; 5(1):1052.

[5] Chen, T., Rui, J., Wang, Q. et al. A mathematical model for simulating the phase-based transmissibility of a novel coronavirus. Infect Dis Poverty 9, 24(2020). https://doi.org/10.1186/s40249-020-00640-3

[6] Carcione José M., Santos Juan E., Bagaini Claudio, Ba Jing. A Simulation of a COVID-19 Epidemic Based on a Deterministic SEIR Model Frontiers in Public Health, 8, 2020, 230 https://www.frontiersin.org/article/10.3389/fpubh.2020.00230

[7] Guanghong, D., Chang, L., Jianqiu, G. et al. SARS epidemical forecast research in mathematical model. Chin.Sci. Bull. 49, 2332–2338 (2004)

[8] WHO, Coronavirus disease, Situation Report – 207. https://www.who.int/emergencies/diseases/novel-coronavirus-2019/situation-reports.

